# Loneliness during strict lockdown: trajectories and predictors during the COVID-19 pandemic in 38,217 adults in the UK

**DOI:** 10.1101/2020.05.29.20116657

**Authors:** Feifei Bu, Andrew Steptoe, Daisy Fancourt

**Author notes:** Corresponding author: Dr Daisy Fancourt, 1-19 Torrington Place, London, WC1E 7HB.

## Abstract

**Rationale:** There are increasing worries that lockdowns and “stay-at-home” orders due to the COVID-19 pandemic could lead to a rise in loneliness, which is recognised as a major public health concern. But profiles of loneliness during the pandemic and risk factors remain unclear.

**Objective:** The current study aimed to examine if and how loneliness levels changed during the strict lockdown and to explore the clustering of loneliness growth trajectories.

**Methods:** Data from 38,217 UK adults in the UCL COVID -19 Social Study (a panel study collecting data weekly during the pandemic) were analysed during the strict lockdown period in the UK (23/03/2020-10/05/2020). The sample was well-stratified and weighted to population proportions of gender, age, ethnicity, education and geographical location. Growth mixture modelling was used to identify the latent classes of loneliness growth trajectories and their predictors.

**Results:** Analyses revealed four classes, with the baseline loneliness level ranging from low to high. In the first a few weeks of lockdown, loneliness levels increased in the highest loneliness group, decreased in the lowest loneliness group, and stayed relatively constant in the middle two groups. Younger adults (*OR*=2.17-6.81), women (*OR*=1.59), people with low income (*OR*=1.3), the economically inactive (*OR*=1.3-2.04) and people with mental health conditions (*OR*=5.32) were more likely to be in highest loneliness class relative to the lowest. Further, living with others or in a rural area, and having more close friends or greater social support were protective.

## Introduction

The COVID-19 pandemic has triggered lockdowns and “stay-at-home” orders in countries around the world. Individuals have been forced to withdraw from usual face-to-face social activities other than with people they live with for substantial periods. This has led to concerns that there could be adverse effects on loneliness, in particular for individuals considered “high risk” for whom stay-at-home orders may be maintained even when orders are officially relaxed for other people (Armitage and Nellums, 2020).

Loneliness is a major public health concern as research has shown associations with heightened risk of mental illness, including depression, generalised anxiety, and suicidal ideation (Beutel et al., 2017). People who are lonely are more likely to develop cardiovascular disease, stroke, and coronary heart disease (Steptoe et al., 2004, Valtorta et al., 2016), as well as experience cognitive decline and develop dementia (Boss et al., 2015, Donovan et al., 2017, Gerst-Emerson and Jayawardhana, 2015, Kuiper et al., 2015). Loneliness is also associated with increased all-cause mortality risk (Lara et al., 2020, Rico-Uribe et al., 2018). Biological studies of the mechanisms underlying these adverse associations have highlighted inflammatory pathways as one explanation for such findings, with loneliness associated with higher levels of inflammation and impaired immune regulation (Cole et al., 2007, Hackett et al., 2012, Walker et al., 2019), which are in turn associated with chronic stress, depression, and cardiovascular risk factors (Dowlati et al., 2010). Behavioural studies of mechanisms have also highlighted the adverse effects of loneliness on health behaviours such as smoking, drinking, and over-eating (Hawkley and Cacioppo, 2003). As such, the potential effects of COVID-19 on loneliness are not just relevant from an individual well-being perspective, but also in terms of the mental and physical health outcomes that could occur as a result.

Recent cross-sectional studies have reported higher levels of loneliness during the COVID-19 pandemic (Bu et al., 2020, Killgore et al., 2020). A longitudinal comparison also shows a significant increase in loneliness among Dutch older adults during the pandemic than late 2019 (van Tilburg et al., 2020). Analyses of Google Trends suggest that Google searches for loneliness increased in the month leading up to lockdowns in Western European countries, with levels then staying high for the fortnight following before returning to usual levels (Brodeur et al., 2020). Another study of American adults examines change in loneliness, finding no significant mean changes in loneliness between January and April, 2020 (Luchetti et al., 2020). It should be noted this study has only three time points with one assessment before the outbreak and two after. This has limited the statistical methodology in depicting the trajectories of loneliness. Also, the longitudinal changes in loneliness may differ across countries where different measures have been taken to tackle the COVID-19 pandemic. It is still unclear what the trajectories of loneliness have been since social distancing or lockdown measures were introduced, and what factors are associated with loneliness trajectories. Cross-sectional data from Spain have suggested that women are at higher risk of experiencing loneliness during the pandemic, as are younger adults, and that higher contact with relatives might be protective (Losada-Baltar et al., 2020), which echoes previous data on usual risk factors (Pinquart and Sörensen, 2003). But much more detailed, longitudinal research is needed.

Loneliness and in particular the identification for factors that could buffer against it, have been highlighted as mental health research priorities in COVID-19 (Holmes et al., 2020). Therefore this study explored trajectories of loneliness during strict lockdown in the UK in a large sample of 35,712 adults tracked across seven weeks from the 23^rd^ March to 10^th^ May, 2020. In particular, the study sought to identify whether loneliness levels changed as the length of lockdown increased, either increasing as individuals became more isolated from others, or decreasing as individuals adapted to circumstances. In other words, if individuals were forced to curtail their usual social activities, how was loneliness affected over the ensuing weeks. It also sought to identify risk and resilience factors for loneliness experiences, including exploring (i) which socio-demographic characteristics or existing mental illness were risk factors for loneliness during lockdown, (ii) whether social factors including living status, social network size and social support protected against experiences of loneliness, and (iii) whether any protective social factors moderated any relationship between mental illness and loneliness.

## Methods

### Participants

We used data from the UCL COVID-19 Social Study; a large panel study of the psychological and social experiences of over 70,000 adults (aged 18+) in the UK during the COVID-19 pandemic. The study commenced on 21^st^ March 2020 (2 days before the lockdown in the UK) involving online weekly data collection from participants for the duration of the COVID-19 pandemic in the UK. Whilst not random, the study has a well-stratified sample that was recruited using three primary approaches. First, snowballing was used, including promoting the study through existing networks and mailing lists (including large databases of adults who had previously consented to be involved in health research across the UK), print and digital media coverage, and social media. Second, more targeted recruitment was undertaken through partnership with recruitment companies focusing on (i) individuals from a low-income background, (ii) individuals with no or few educational qualifications, and (iii) individuals who were unemployed. Third, the study was promoted via partnerships with third sector organisations to vulnerable groups, including adults with pre-existing mental illness, older adults, and carers. Full detail on the recruitment and sampling is available in the study User Guide (www.covidsocialstudy.org). The study was approved by the UCL Research Ethics Committee [12467/005] and all participants gave informed consent. In this study, we focused on participants who had at least three repeated measures between 23^rd^ March and 10^th^ May 2020. This provided us with data from 42,411 participants. The data were analysed using a complete case analysis approach where 10% participants with missing data were excluded, providing a final analytic sample size of 38,217.

### Measures

Loneliness was measured using the three-item UCLA loneliness scale (UCLA-3). The questions include: 1) how often do you feel lack companionship? 2) how often do you feel isolated from others? 3) how often do you feel left out? Responses to each question were scored on a three-point Likert scale ranging from hardly ever/never, to some of the time, to often. Using the sum score, this provided a loneliness scale ranging from 3 to 9, with a higher score indicating increased loneliness.

Covariates included age groups (18-29, 30-45, 46-59 and 60+), gender (woman vs. man), ethnicity (non-white vs. white), education (low: GCSE or below, medium: A levels or equivalent, high: degree or above), low income (household annual income <£30,000 vs. higher household annual income), employment status (employed, unemployed, student and inactive other), and area of living (rural vs. urban). Given living status (alone vs with others) is highly correlated with marital status, meaning that only one of these factors could be included within the model to avoid multicollinearity, we included living status as it is a clearer indicator of social interactions in the home.

We also assessed social relationship measures, including having large friend network (number of close friends ≥3), high usual social contact (at least weekly face-to-face contact) and high perceived social support (measured using the brief form of the perceived social support (F-SozU K-6) scale (Lin et al., 2019). Each item of F-SozU K-6 is rated on a five-point scale from “not true at all” to “very true”, with higher scores indicating higher levels of perceived social support. Minor adaptations were made to the language in the scale to make it relevant to experiences during COVID-19 (see Supplementary Table S1 for a comparison of changes). In addition, we also examined mental illness as a predictor of loneliness trajectories, through participant report of clinical diagnoses of depression, anxiety, or other psychiatric conditions (yes/no).

### Analysis

To identify growth trajectories of loneliness and their predictors, we used the growth mixture modelling (GMM) approach. The conventional growth modelling approach assumes one homogeneous growth trajectory, allowing individual growth factors to vary randomly around the overall mean. GMM, on the other hand, relaxes this assumption and enables researchers to explore distinctive latent growth trajectory classes. For detailed explanation of GMM, refer to Muthén and Asporouhov (2006).

The model specification in this study is presented in Figure 1. The seven repeated measures of loneliness were used as the indicators of the latent growth factors, the intercept and slope, which were influenced by the latent growth trajectory class. In this model, we made no assumption about the shape of growth trajectories which was left to be determined by the data. This was achieved by setting the time scores as free parameters (*), except for two fixed to 0 and 1 for the model to be identified. Moreover, the residuals of adjacent loneliness measures were specified to be correlated to capture the possibility of unknown shared causes of the covariance between repeated measures.

**Figure 1.**
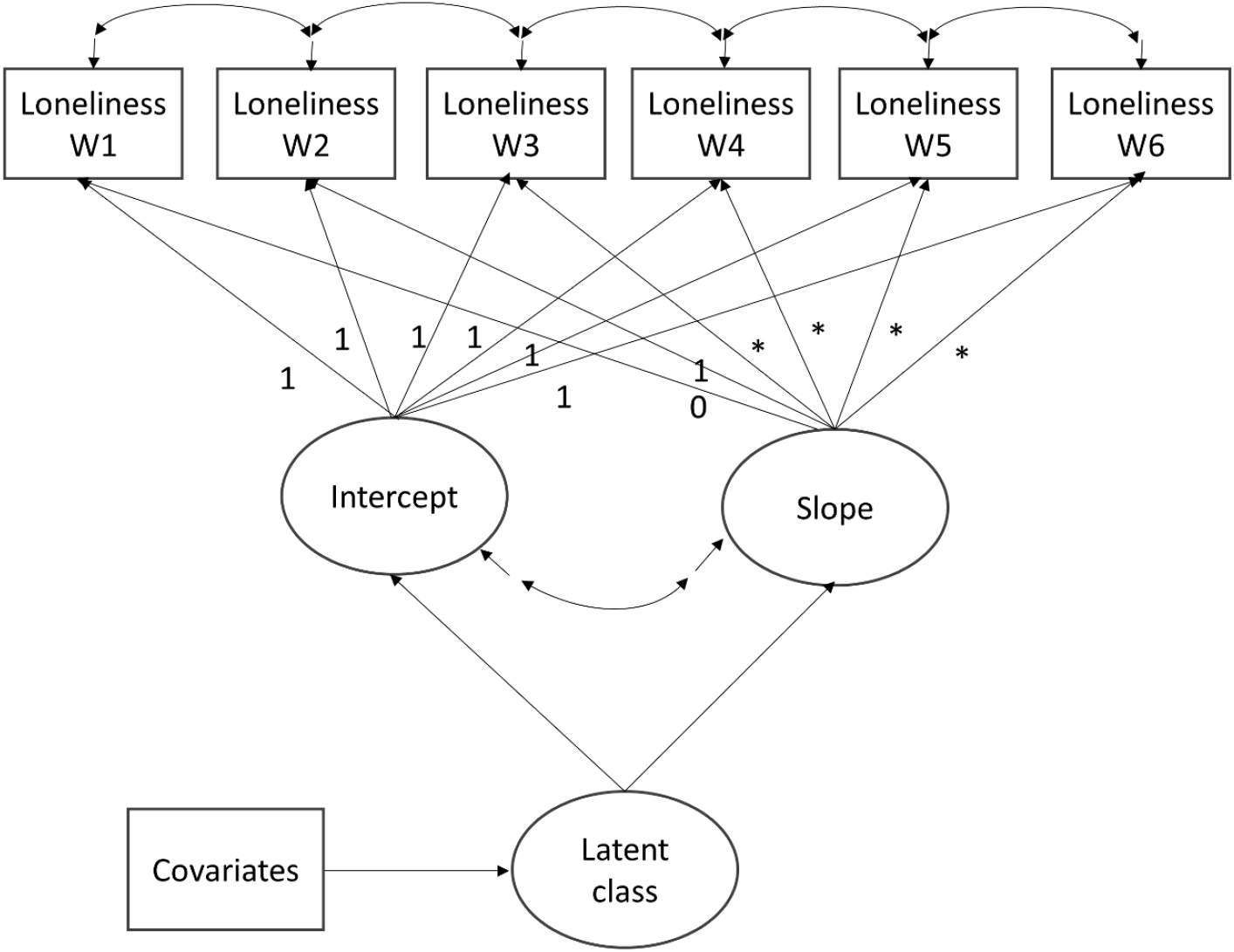
Overall growth mixture model specification

Starting with the unconditional GMM, we compared models with different number of classes on the basis of Bayesian criteria, Bayesian information criterion (BIC) and sample-size adjusted Bayesian information criterion (ABIC), along with Vuong-Lo-Mendell-Rubin likelihood ratio (LMR-LR) test and Adjusted Lo-Mendell-Rubin likelihood ratio (ALMR-LR) test. After identifying the optimal number of classes, we introduced covariates to explain the observed heterogeneity between classes.

Weights were applied throughout the analyses. All data were weighted to the proportions of gender, age, ethnicity, education and country of living obtained from the Office for National Statistics (ONS). The descriptive and regression analyses were implemented in Stata v15 and GMM in Mplus Version 8.

## Results

### Trajectories of loneliness during lockdown

The first step was to determine the optimal number of latent trajectory classes. The model fit indices of models under comparison are presented in Table 1. Across models with different number of classes, the 4-class model had the lowest BIC and ABIC. In addition, the LMR-LR and ALMR-LR tests in the 5-class model both had a P-value>0.05, confirming that the 4-class model was favoured.

**Table 1.** Model fit indices for different model specifications

| Model specification | Parameters | BIC | ABIC | LMR-LR | ALMR-LR | Entropy |
| --- | --- | --- | --- | --- | --- | --- |
| 1-class GMM | 23 | 594074 | 594001 | NA | NA | NA |
| 2-class GMM | 26 | 586583 | 586500 | <0.001 | <0.001 | 0.771 |
| 3-class GMM | 29 | 574046 | 573953 | <0.001 | <0.001 | 0.922 |
| **4-class GMM** | **32** | **570160** | **570058** | **<0.001** | **<0.001** | **0.883** |
| 5-class GMM | 35 | 569129 | 569018 | 0.164 | 0.174 | 0.889 |

The estimated growth trajectory for each class is shown in Figure 2. Generally speaking, loneliness was stable over the seven weeks of strict lockdown. However, for the class with the highest initial status (LC4 the loneliest, 14.3%), there was about one point increase in loneliness from week 1 to 5. This was followed by a decrease in week 6, stabilising again in week 7. Changes in loneliness across time were also observed in the lowest loneliness class (LC1, 48.2%), with loneliness decreasing in the first five weeks before rebounding in week 6. But there was no sign of continuing increase in week 7.

**Figure 2.**
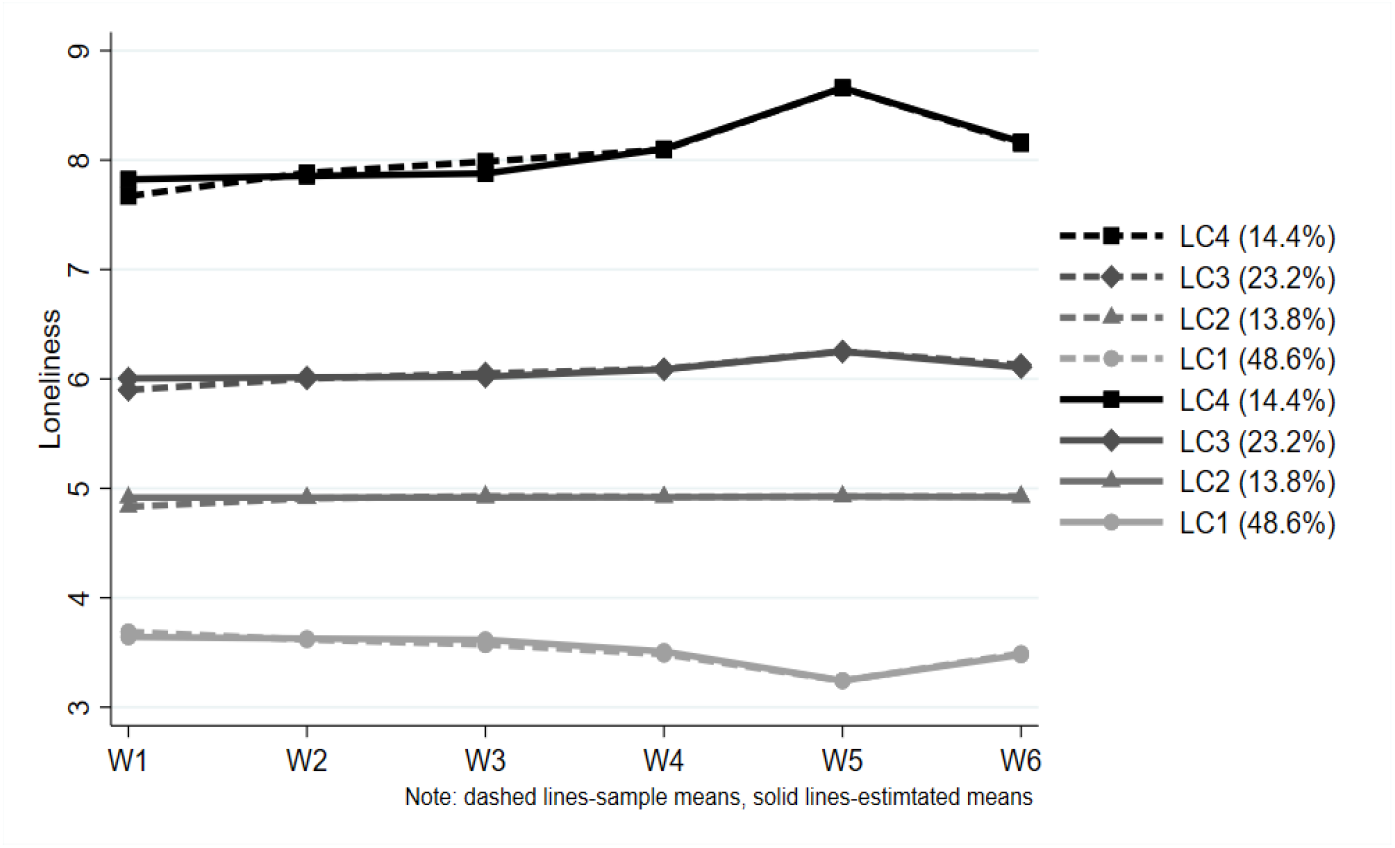
Estimated growth trajectory for each latent class based on the 4-class unconditional GMM with free time scores

### Protective factors for experiences of loneliness during lockdown

Table 2 presents the results from the GMM with covariates. Using LC1 (lowest loneliness) as the reference, the odds of being in a higher loneliness class were higher in a dose-response pattern with age. Adults aged 18-29 had a 6 times higher odds of being in the highest-risk class compared to adults aged 60+, while adults aged 30-45 had a 4 times higher odds and adults aged 46-59 had a 2.1 times higher odds. Women had a higher odds of being in a higher loneliness class, with a 71% higher odds of being in the loneliest class.

**Table 2.** Estimated odds ratios, standard errors, *p* values of the predictors of latent growth trajectory classes (*N*=53,712)

|  |  | Med-low (vs. lowest) |  |  | Med-high (vs. lowest) |  |  | Highest (vs. lowest) |  |  |
| --- | --- | --- | --- | --- | --- | --- | --- | --- | --- | --- |
|  |  | LC2 (vs. LC1) |  |  | LC3 (vs. LC1) |  |  | LC4 (vs. LC1) |  |  |
| Variables |  | *OR* | *SE* | *p* | *OR* | *SE* | *p* | *OR* | *SE* | *p* |
| Age | 18-29 | 2.58 | 0.43 | 0.000 | 3.33 | 0.44 | 0.000 | 6.03 | 0.97 | 0.000 |
|  | 30-45 | 1.54 | 0.18 | 0.003 | 2.47 | 0.21 | 0.000 | 4.00 | 0.45 | 0.000 |
|  | 46-59 | 1.11 | 0.11 | 0.319 | 1.66 | 0.13 | 0.000 | 2.08 | 0.21 | 0.000 |
|  | 60+ | Ref. | Ref. | Ref. | Ref. | Ref. | Ref. | Ref. | Ref. | Ref. |
| Gender | Women (Ref. men) | 1.37 | 0.11 | 0.001 | 1.49 | 0.09 | 0.000 | 1.71 | 0.14 | 0.000 |
| Ethnicity | Non-white (Ref. white) | 1.28 | 0.23 | 0.228 | 1.06 | 0.14 | 0.656 | 0.96 | 0.16 | 0.810 |
| Education | GCSE or below | Ref. | Ref. | Ref. | Ref. | Ref. | Ref. | Ref. | Ref. | Ref. |
|  | A-levels or equivalent | 1.06 | 0.11 | 0.571 | 1.11 | 0.09 | 0.210 | 1.09 | 0.11 | 0.432 |
|  | Degree or above | 1.00 | 0.10 | 0.993 | 1.02 | 0.08 | 0.838 | 0.86 | 0.09 | 0.117 |
| Household income | Low (<30k) (Ref. high) | 1.00 | 0.09 | 0.976 | 1.23 | 0.08 | 0.006 | 1.33 | 0.12 | 0.006 |
| Employment status | Employed | Ref. | Ref. | Ref. | Ref. | Ref. | Ref. | Ref. | Ref. | Ref. |
|  | Unemployed | 1.26 | 0.30 | 0.388 | 1.33 | 0.30 | 0.261 | 1.75 | 0.41 | 0.066 |
|  | Student | 1.36 | 0.36 | 0.318 | 1.44 | 0.29 | 0.125 | 2.14 | 0.48 | 0.017 |
|  | Inactive other | 0.98 | 0.10 | 0.793 | 0.93 | 0.07 | 0.338 | 1.23 | 0.11 | 0.043 |
| Mental health | Diagnosed condition | 1.90 | 0.26 | 0.000 | 2.87 | 0.28 | 0.000 | 5.00 | 0.55 | 0.000 |
| Living status | Living with others (Ref. alone) | 0.45 | 0.04 | 0.000 | 0.43 | 0.03 | 0.000 | 0.25 | 0.02 | 0.000 |
| Area of living | Rural (Ref. urban) | 0.92 | 0.08 | 0.288 | 0.86 | 0.05 | 0.006 | 0.76 | 0.07 | 0.000 |
| Number of close friends | Large ($\geq 3$) (Ref. small) | 0.99 | 0.09 | 0.900 | 0.83 | 0.06 | 0.003 | 0.58 | 0.05 | 0.000 |
| Usual face-to-face contact | At least weekly (Ref. < weekly) | 0.93 | 0.08 | 0.370 | 1.16 | 0.07 | 0.027 | 1.12 | 0.09 | 0.178 |
| Perceived social support | High (sum score $\geq 18$) (Ref. low) | 0.54 | 0.05 | 0.000 | 0.25 | 0.02 | 0.000 | 0.11 | 0.01 | 0.000 |

Ethnicity was not a risk factor for being lonelier, and nor was education. Low household income did not predict membership of the LC2 class (medium-low loneliness) but did predict membership of higher loneliness classes, with people earning less than £30,000 per year having a 33% higher odds of being in the highest loneliness class. Relative to people who were employed, being unemployed was not a risk factor for being lonely, but students and people who were inactive (e.g. home makers or people who were retired) had 2.1 and 1.2 times the odds of being in the highest loneliness class respectively. Mental health was a significant predictor of higher loneliness, with people with a diagnosed mental health condition having a 5 times higher odds of being in the highest loneliness class.

In looking at which social factors might be protective, living with others was protective against loneliness, with a 75% lower odds of being in the highest risk class compared to people living alone. Living in a rural area was also protective, with a 24% lower odds of being in the highest loneliness class. People with a larger circle of close friends had a 42% lower odds of being in the highest loneliness class, while people with low perceived social support had an 89% lower odds of being in the highest loneliness class.

However, when looking at interactions, there was no evidence that any social factors moderated the relationship between mental illness and loneliness (see Supplementary Table S3).

## Discussion

This is the first study to examine the growth trajectories and predictors of loneliness during lockdown due to the Covid-19 pandemic. We identified four major classes of loneliness, ranging from low to high. In the seven weeks of strict lockdown, loneliness levels increased slightly in the highest loneliness group, decreased slightly in the lowest loneliness group, and stayed relatively constant in the middle two groups. But there was some regression to the mean in the highest and lowest groups into week six. Demographic factors such as younger age, being female, low household income, and being a student were all risk factors for being in a higher loneliness class, as was a diagnosis of a mental health condition. Living with others, living in a rural area, having more close friends, and having greater perceived social support were all protective against higher loneliness levels, even during lockdown when usual face-to-face contact was disrupted. There was only limited evidence that loneliness was higher for people who usually had more face-to-face contact, and this did not predict being in the highest loneliness class. However, there was no evidence that protective social factors moderated the relationship between poor mental health and risk of loneliness.

It is concerning that 14% of participants were in the highest loneliness class, with average loneliness levels of around 8-8.5. Data on national levels of loneliness in the UK outside of COVID-19 suggest that usually only 6.1% of adults experience scores of 8 or 9 on the UCLA-3 loneliness scale (Bu et al., 2020). The UCL COVID-19 Social Study did not use a random sample, so we do not claim prevalence figures, but the findings nonetheless suggest that there are a substantial number of people feeling high levels of loneliness. It is also notable that loneliness for this group increased, particularly around the fifth week of the lockdown. This period coincided with two weeks following the Easter bank holiday, a traditional moment of national celebration, so may have been a response to being unable to engage in planned social activities. But it is also possible that fatigue relating to lockdown may have exacerbated existing loneliness symptoms. For the lowest loneliness class, these findings were the opposite, with a gradual decrease over the first month of lockdown, with lowest levels recorded in week five. Whether the indications of return to levels prior to the week-five change since in the highest and lowest loneliness class is a result of any change in announcements around the easing of lockdown or simply a regression to the mean remains to be explored in future analyses. Nevertheless, it is striking that there were no marked changes over time, and that loneliness levels appear to have been established early in the lockdown period. This suggests that whilst the curtailing of social activities is associated with higher than usual levels of loneliness, there is little evidence either of adaptation of loneliness responses to the circumstances, nor growing sensations of loneliness. However, as this study explored a highly specific social situation which, importantly, affected people globally, there was little opportunity for feelings such as fear of missing out, which may play a role in experiences of loneliness (Baker et al., 2016).

The findings on women and young people being highest risk for loneliness echoes previous research on risk factors both during the pandemic (Losada-Baltar et al., 2020), and ordinarily (Pinquart and Sörensen, 2003). Young people may engage in more gregarious social activity in normal life, so suffer more during isolation. Similarly, our finding that people with a diagnosed mental illness had a higher odds of being lonelier aligns with research showing a bidirectional link between loneliness and mental health (Wang et al., 2018). A number of social factors, though, were identified as resilience factors that protected against loneliness, including living with others, living in a rural location, having three or more close friends, and having high perceived social support. This echoes previous research suggesting a relationship between social network size and social support and lower risk of loneliness (Cacioppo et al., 2009, Cacioppo and Cacioppo, 2014, Domènech-Abella et al., 2017, Lee and Goldstein, 2016). It is notable that usually having frequent face-to-face contact was not a risk factor for higher loneliness during lockdown, which suggests that experiencing a sudden change in social behaviours does not in itself predict loneliness.

It is important to consider how to tackle loneliness during the COVID-19 pandemic. The results presented here highlight the exceptionally high level of loneliness among younger adults, especially those aged 18-29 and students. There have been calls for the promotion of digital technologies to bridge social distance, as well as the development of outreach and screening for loneliness alongside associated mental health conditions so that social support can be provided (Galea et al., 2020). Our results suggest that these may be supportive given the finding that perceived social support is protective against loneliness. However, it is notable that none of the protective social factors moderated the relationship between mental illness and loneliness. Previous work has suggested that social factors can mediate the relationship between loneliness and depression (Liu et al., 2016), and much research has shown the relationship between social factors, loneliness and trajectories of depression (Van Den Brink et al., 2018). But less work has been done looking at how mental health and social factors interact to predict trajectories of loneliness. Some previous research has suggested that factors such as social support do not consistently buffer the relationship between loneliness and stress (Lee and Goldstein, 2016), with context playing a key role. This raises the question as to whether the social situation during COVID-19 provides a unique context for understanding the interplay between loneliness and mental health. The pandemic has disrupted usual social behaviours and is posing a major challenge for mental health. As such, having more friends may be protective against loneliness to a certain extent amongst individuals without diagnosed mental health conditions, but may be insufficient in the face of higher levels of anxiety or depression. Indeed, it is notable that loneliness during COVID-19 has been associated with poorer mental health and greater worries around the health impact of the virus (Cerami et al., 2020, Okruszek et al., 2020), suggesting that there may be an exacerbation of worries amongst those who are lonely. Therefore, strategies to address loneliness in people with mental illness may require greater nuance than merely providing extra social support. Schemes that have previously been used to address loneliness in individuals with mental illness, such as social prescribing schemes that combine social activities with clinical support, may be promising avenues to pursue. Further, addressing loneliness may be an important target in reducing symptoms of anxiety and depression in individuals with mental illness (Mann et al., 2017).

## Limitations

A main limitation of this study is that the UCL Covid-19 Social Study did not use a random sample, and therefore our reported statistics cannot be taken as accurate prevalence for loneliness in the UK. The study does have a large sample size with wide heterogeneity, including good stratification across all major socio-demographic groups, and analyses were weighted on the basis of population estimates of core demographics, with the weighted data showing good alignment with national population statistics and another large scale nationally representative social survey. But we cannot rule out the possibility that the study inadvertently attracted individuals experiencing more extreme psychological experiences, with subsequent weighting for demographic factors failing to fully compensate for these differences. Further, these analyses focused on trajectories during lockdown, but how this compares to individuals’ usual experiences of loneliness remains to be explored in future studies.

## Conclusions

Overall, these findings suggest that perceived levels of loneliness in the seven weeks of strict lockdown during COVID-19 were relatively stable in the UK, but for many people these levels were high with no signs of improvement. People with pre-existing mental health diagnoses, younger adults, women and students were at greatest risk of experiencing high levels of loneliness, but certain social factors such as living with others, having close friends, and having strong perceived social support were protective. There is currently much policy interest in trying to address loneliness within society, both within the context of COVID-19 and more generally. Our results highlight which groups are at a higher risk and suggest that more interventions and guidelines are needed to help reduce loneliness.

## Data Availability

Anonymous data will be made available following the end of the pandemic.

## Acknowledgement

This Covid-19 Social Study was funded by the Nuffield Foundation [WEL/FR-000022583], but the views expressed are those of the authors and not necessarily the Foundation. The study was also supported by the MARCH Mental Health Network funded by the Cross-Disciplinary Mental Health Network Plus initiative supported by UK Research and Innovation [ES/S002588/1], and by the Wellcome Trust [221400/Z/20/Z]. DF was funded by the Wellcome Trust [205407/Z/16/Z]. The researchers are grateful for the support of a number of organisations with their recruitment efforts including: the UKRI Mental Health Networks, Find Out Now, UCL BioResource, HealthWise Wales, SEO Works, FieldworkHub, and Optimal Workshop. The funders had no final role in the study design; in the collection, analysis and interpretation of data; in the writing of the report; or in the decision to submit the paper for publication. All researchers listed as authors are independent from the funders and all final decisions about the research were taken by the investigators and were unrestricted. All authors had full access to all of the data (including statistical reports and tables) in the study and can take responsibility for the integrity of the data and the accuracy of the data analysis.

## Supplementary Material

**Table S1:** Comparison of items in the original and revised Perceived Social Support Questionnaire (F-SozU K-6).

| Original | Adapted for COVID-19<br>In the past week, I feel... |
| --- | --- |
| I experience a lot of understanding and security from others | I have experienced a lot of understanding and support from others |
| I know a very close person whose help I can always count on | I have a very close person whose help I can always count on |
| If necessary, I can easily borrow something I might need from neighbours or friends | If necessary, I can easily borrow something I need from neighbours or friends |
| I know several people with whom I like to do things | I have people with whom I can spend time and do things together |
| When I am sick, I can without hesitation ask friends and family to take care of important matters for me | If I get sick, I have friends and family who will take care of me |
| If I am down, I know to whom I can go without hesitation | If I am feeling down, I have people I can talk to without hesitation |

**Table S2.** Descriptive statistic of the explanatory variables (*N*=38,217, weighted)

| Variables |  | Percentages |
| --- | --- | --- |
| Age | 18-29 | 19.50% |
|  | 30-45 | 26.10% |
|  | 46-59 | 24.10% |
|  | 60+ | 30.30% |
| Gender | Women (vs. men) | 50.60% |
| Ethnicity | Non-white (vs. white) | 12.80% |
| Education | GCSE or below | 32.70% |
|  | A-levels or equivalent | 33.90% |
|  | Degree or above | 33.41% |
| Household income | Low (<30k) (vs. high) | 47.60% |
| Employment status | Employed | 57.68% |
|  | Unemployed | 2.63% |
|  | Student | 7.97% |
|  | Inactive other | 31.71% |
| Mental health | Diagnosed condition | 19.86% |
| Living status | Living with others (vs. alone) | 81.33% |
| Area of living | Rural (vs. urban) | 20.49% |
| Number of close friends | Large ($\geq 3$) (vs. small) | 69.61% |
| Usual face-to-face contact | At least weekly (vs. < weekly) | 67.78% |
| Perceived social support | High (sum score $\geq 18$) (vs. low) | 73.28% |

**Table S3.**
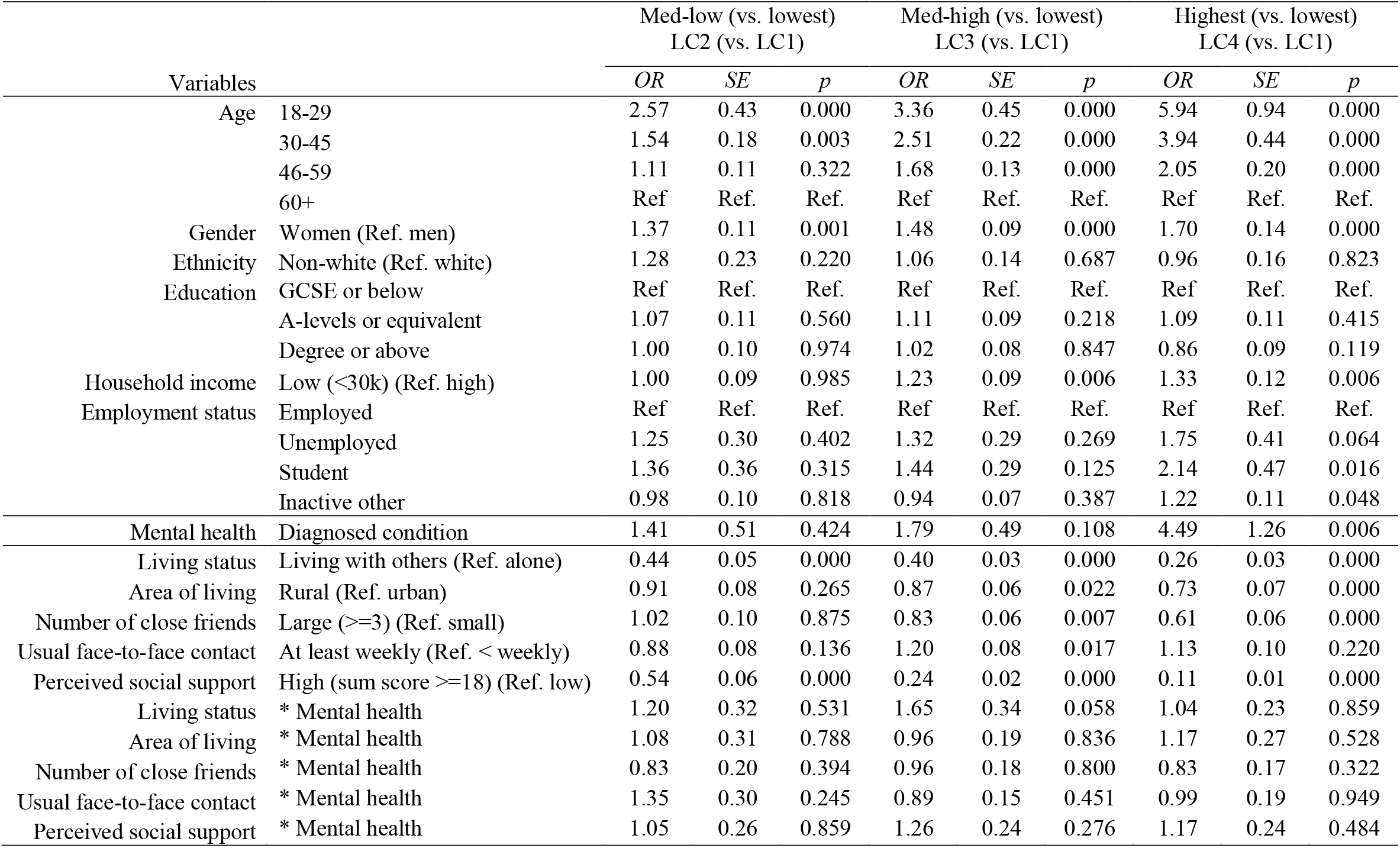
Estimated odds ratios, standard errors, *p* values of the predictors of latent growth trajectory classes, with interaction terms (*N*=38,217)

|  |  | Med-low (vs. lowest)<br>LC2 (vs. LC1) |  |  | Med-high (vs. lowest)<br>LC3 (vs. LC1) |  |  | Highest (vs. lowest)<br>LC4 (vs. LC1) |  |  |
| --- | --- | --- | --- | --- | --- | --- | --- | --- | --- | --- |
| Variables |  | *OR* | *SE* | *p* | *OR* | *SE* | *p* | *OR* | *SE* | *p* |
| Age | 18-29 | 2.57 | 0.43 | 0.000 | 3.36 | 0.45 | 0.000 | 5.94 | 0.94 | 0.000 |
|  | 30-45 | 1.54 | 0.18 | 0.003 | 2.51 | 0.22 | 0.000 | 3.94 | 0.44 | 0.000 |
|  | 46-59 | 1.11 | 0.11 | 0.322 | 1.68 | 0.13 | 0.000 | 2.05 | 0.20 | 0.000 |
|  | 60+ | Ref. | Ref. | Ref. | Ref. | Ref. | Ref. | Ref. | Ref. | Ref. |
| Gender | Women (Ref. men) | 1.37 | 0.11 | 0.001 | 1.48 | 0.09 | 0.000 | 1.70 | 0.14 | 0.000 |
| Ethnicity | Non-white (Ref. white) | 1.28 | 0.23 | 0.220 | 1.06 | 0.14 | 0.687 | 0.96 | 0.16 | 0.823 |
| Education | GCSE or below | Ref. | Ref. | Ref. | Ref. | Ref. | Ref. | Ref. | Ref. | Ref. |
|  | A-levels or equivalent | 1.07 | 0.11 | 0.560 | 1.11 | 0.09 | 0.218 | 1.09 | 0.11 | 0.415 |
|  | Degree or above | 1.00 | 0.10 | 0.974 | 1.02 | 0.08 | 0.847 | 0.86 | 0.09 | 0.119 |
| Household income | Low (<30k) (Ref. high) | 1.00 | 0.09 | 0.985 | 1.23 | 0.09 | 0.006 | 1.33 | 0.12 | 0.006 |
| Employment status | Employed | Ref. | Ref. | Ref. | Ref. | Ref. | Ref. | Ref. | Ref. | Ref. |
|  | Unemployed | 1.25 | 0.30 | 0.402 | 1.32 | 0.29 | 0.269 | 1.75 | 0.41 | 0.064 |
|  | Student | 1.36 | 0.36 | 0.315 | 1.44 | 0.29 | 0.125 | 2.14 | 0.47 | 0.016 |
|  | Inactive other | 0.98 | 0.10 | 0.818 | 0.94 | 0.07 | 0.387 | 1.22 | 0.11 | 0.048 |
| Mental health | Diagnosed condition | 1.41 | 0.51 | 0.424 | 1.79 | 0.49 | 0.108 | 4.49 | 1.26 | 0.006 |
| Living status | Living with others (Ref. alone) | 0.44 | 0.05 | 0.000 | 0.40 | 0.03 | 0.000 | 0.26 | 0.03 | 0.000 |
| Area of living | Rural (Ref. urban) | 0.91 | 0.08 | 0.265 | 0.87 | 0.06 | 0.022 | 0.73 | 0.07 | 0.000 |
| Number of close friends | Large (>=3) (Ref. small) | 1.02 | 0.10 | 0.875 | 0.83 | 0.06 | 0.007 | 0.61 | 0.06 | 0.000 |
| Usual face-to-face contact | At least weekly (Ref. < weekly) | 0.88 | 0.08 | 0.136 | 1.20 | 0.08 | 0.017 | 1.13 | 0.10 | 0.220 |
| Perceived social support | High (sum score >=18) (Ref. low) | 0.54 | 0.06 | 0.000 | 0.24 | 0.02 | 0.000 | 0.11 | 0.01 | 0.000 |
| Living status | * Mental health | 1.20 | 0.32 | 0.531 | 1.65 | 0.34 | 0.058 | 1.04 | 0.23 | 0.859 |
| Area of living | * Mental health | 1.08 | 0.31 | 0.788 | 0.96 | 0.19 | 0.836 | 1.17 | 0.27 | 0.528 |
| Number of close friends | * Mental health | 0.83 | 0.20 | 0.394 | 0.96 | 0.18 | 0.800 | 0.83 | 0.17 | 0.322 |
| Usual face-to-face contact | * Mental health | 1.35 | 0.30 | 0.245 | 0.89 | 0.15 | 0.451 | 0.99 | 0.19 | 0.949 |
| Perceived social support | * Mental health | 1.05 | 0.26 | 0.859 | 1.26 | 0.24 | 0.276 | 1.17 | 0.24 | 0.484 |

**Figure S1.**
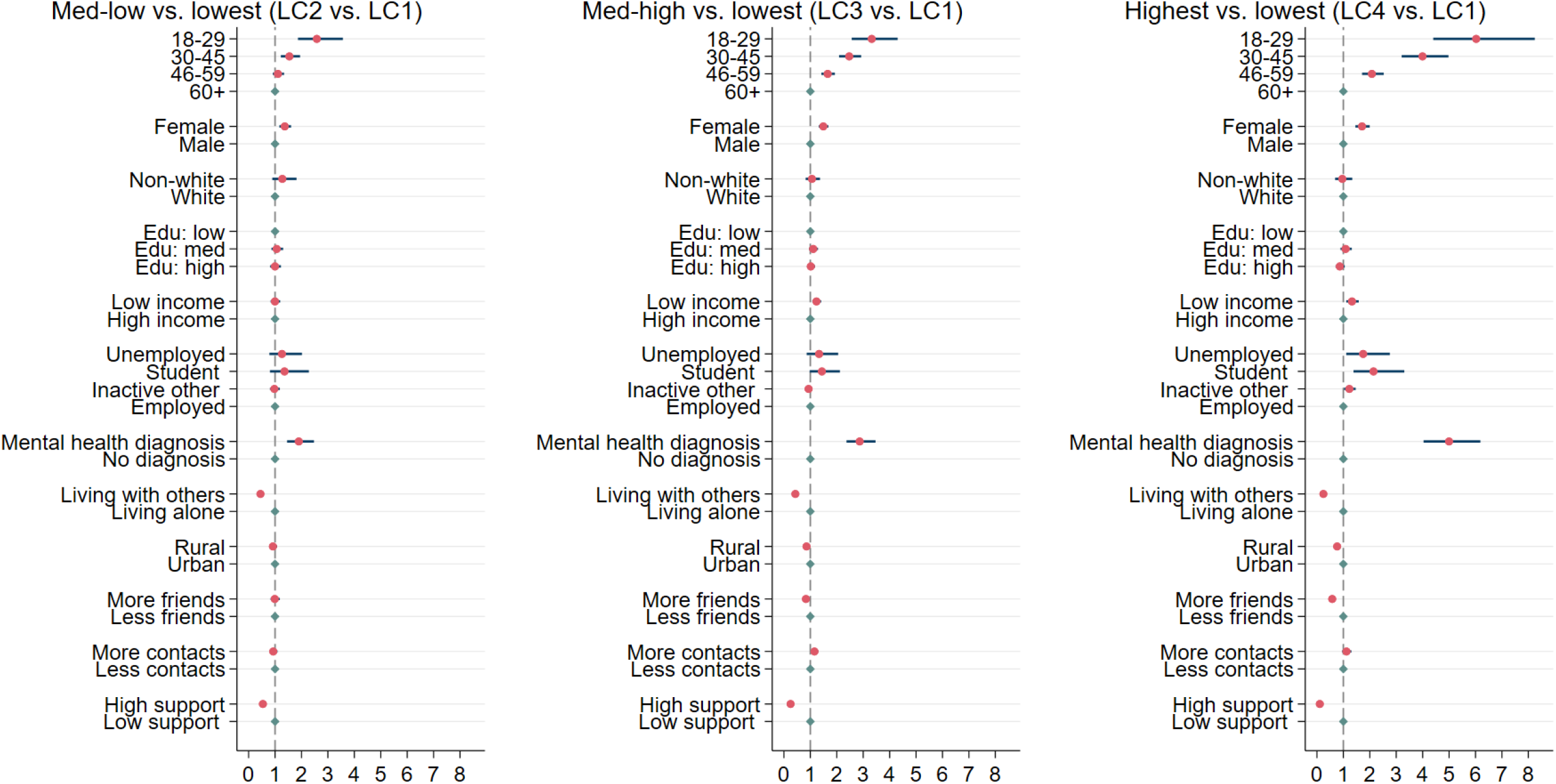
Estimated odds ratios and 95% confidence intervals for the predictors of latent growth trajectory classes (Table 2)

